# Impact of Prior Dengue Infection on the Severity and Outcomes of Subsequent Infections: A Meta-Analysis of the Placebo Arm of Clinical Trials

**DOI:** 10.1101/2024.06.06.24308498

**Authors:** Alejandro Macchia, Silvana Figar, Cristián Biscayart, Fernán González Bernaldo de Quirós

## Abstract

**Importance:** The increasing incidence and prevalence of dengue in Latin America is well-documented. Historical case-control series also highlight that secondary infections are a risk factor for severe dengue, hospitalization, and death. This has generated alarm among some sectors of the population and the scientific community. However, there has been no examination based on the results of randomized clinical trials that analyzes the risk of severe events in individuals with and without prior dengue infection.

**Objective:** To evaluate the association between serologically confirmed prior dengue infection (DVC) and the subsequent risk of DVC, severe dengue (DS), dengue hospitalization (DHOSP), dengue-related death, and all-cause mortality.

**Methods:** A systematic review and meta-analysis were conducted following PRISMA guidelines. Studies were searched in PubMed, CINAHL, Medline, Cochrane Library, and Web of Science, including only phase III randomized clinical trials of vaccine efficacy with data on participants in the placebo groups and information on previous infections. Random-effects models were applied to calculate combined odds ratios (OR), and heterogeneity among studies was assessed.

**Results:** A total of four studies corresponding to three phase III clinical trials were included. Participants with prior infection had a lower likelihood of developing DVC during follow-up (OR: 0.85; 95% CI: [0.75; 0.98]; p=0.024) and the same risk of dengue hospitalization as those without prior dengue (OR: 1.18; 95% CI: [0.92; 1.53]; p=0.198). However, they had a higher rate of DS during follow-up (OR: 2.91; 95% CI: [1.23; 6.87]; p=0.015). No dengue-related deaths were observed in any of the clinical trials during follow-up.

**Conclusions:** Prior dengue infection significantly reduces the risk of DVC and increases the risk of DS, although it does not significantly affect the risk of dengue hospitalization or dengue-related death during follow-up. The findings of this study highlight the need to reconsider the value of prior infection as an independent risk factor.

## Introduction

In the context of rising incidence and prevalence of dengue (1,2), several studies have analyzed factors related to severe infections (3). Among these factors, secondary infection has been identified as a significant risk factor for severe dengue (DS) in some studies. A review (3) examining 22 studies found that secondary infections were associated with a significantly higher risk of DS (OR [95% CI]: 2.69 [2.08-3.48]). These results are materially similar to another systematic review of observational studies that found an associated risk of 1.75 [1.26-2.42] (4). However, a prospective study conducted in Peru (5) investigating the role of secondary infections in the context of the American genotype of dengue virus (DEN-2) concluded that, despite the high prevalence of secondary infections, they were not associated with an increased risk of DS. This suggests that the American genotype of DEN-2 might not have the characteristics necessary to induce severe forms of the disease. Another study (6) examined how previous infections with different DENV serotypes affect the risk and severity of subsequent infections, finding that the presence of pre-existing heterotypic antibodies significantly reduces the risk of DS in subsequent infections (6).

Although narrative reviews (7) emphasize secondary infection as a predisposing factor for DS and dengue hospitalization (DHOSP), not all reviews are consistent (3–6). The reasons for this disagreement are numerous, but inherent biases in using cohorts from uncontrolled studies and recall bias in severe patients may contribute.

The emergence of randomized clinical trials (RCTs) examining the efficacy of various vaccines for the prevention of virologically confirmed dengue (DVC), DS, DHOSP, and dengue-related death provides an opportunity to study this topic with a higher level of evidence. Unlike retrospective case-control studies, clinical trials have close prospective surveillance and documentation of events, making them less prone to observation bias.

This study conducted a systematic review and meta-analysis to investigate the role of secondary infections in the occurrence of DVC, DS, DHOSP, and dengue-related death among patients randomized to the placebo/control arm of phase III clinical trials, aiming to determine the association of prior infection with severe forms of dengue.

## Materials and Methods

### Study Design

The PRISMA guidelines were used to guide this systematic review. This review involved several stages: defining keywords, searching databases for article selection, critical evaluation of studies, data selection and analysis, and presentation and interpretation of results. The research protocol is registered with PROSPERO under registration number 542370.

### Inclusion Criteria

Inclusion criteria included peer-reviewed original articles written in English and studies that examined the immunogenicity, safety, and efficacy of various dengue vaccines, without limitations on the years of publication. Only the final studies for each vaccine were reported to eliminate duplications. Only studies that reported the number of participants with documented prior infection in the placebo/control arm were included.

### Exclusion Criteria

Exclusion criteria included articles not written in English and any study that was not a phase III randomized clinical trial or lacked a control or placebo group.

### Search Strategy

Article search strategies were conducted in PubMed, CINAHL, Medline, Cochrane Library, and Web of Science from January 1994 to March 2024 (supplement 1).

Additionally, manual searches (including scanning reference lists) were conducted to identify articles that might not have been included in the initial search strategy.

Two independent investigators (AM, FQ) assessed the title and abstract of all articles according to eligibility criteria for population, intervention, comparison, and study design. The full text of all potentially eligible studies was obtained, and two investigators (AM, FQ) assessed their eligibility. Any disagreements were resolved through discussion. Reasons for exclusion of clinical trials and the selection process were recorded in the PRISMA flow diagram.

### Risk of Bias

The risk of bias in each randomized trial was assessed using the Risk of Bias 2 (RoB 2) tool developed by the Cochrane Collaboration. The five domains of bias considered in this tool were: bias arising from the randomization process, bias due to deviations from intended interventions, bias due to missing outcome data, bias in outcome measurement, and bias in the selection of the reported result.

### Rate of overall clinical events during follow-up

To estimate the risk and 95% confidence intervals (CI) of dengue infections per 100,000 person-years for each study, we collected data from each different clinical trial. For each study, we recorded the total number of participants, dengue infections, severe dengue, hospitalizations for dengue, all-cause deaths and dengue related deaths. We collected and use each study follow-up period in years. We calculated the risk of each of these outcomes per 100,000 person-years for each study. The risk was computed by dividing the total number of infections by the product of the total number of participants and the follow-up period, then multiplying by 100,000. We computed the probability of each single outcome by dividing the total number of events by the product of the total number of participants and the follow-up period. The standard error (SE) was calculated using the formula for the standard error of a proportion. The 95% confidence intervals for the risk were then determined by subtracting and adding 1.96 times the standard error to the probability of every outcome and multiplying the results by 100,000. After estimating the risk and 95% confidence intervals (CI) for each individual study, we proceeded to calculate the overall weighted risk across all studies. To obtain the overall risk, we calculated the weights for each study based on the inverse of the variance (standard error squared) of their respective risk estimates. The overall weighted risk was then computed by summing the products of each study’s weight and risk and dividing by the sum of the weights. The global standard error was derived by taking the square root of the inverse of the sum of the weights.

### Calculation of Association Between Prior Infection and Outcomes of Interest

Separate meta-analyses were conducted for each of the outcomes of interest, identified as DVC, DS, DHOSP, overall mortality and dengue-related death, using the inverse variance random-effects method to calculate the combined odds ratio (OR) for each included study. Each meta-analysis is presented through a forest plot, displaying the central estimates along with their 95% confidence intervals (CI). To assess heterogeneity among the studies included in each meta-analysis, the I² statistic was used. The I² statistic measures the percentage of total variation across studies that is due to heterogeneity rather than chance. Substantial heterogeneity was considered when the I² value exceeded 40% and the p-value associated with the chi-squared (X²) test was less than 0.10. The identification of substantial heterogeneity also influenced the decision to use a random-effects model for calculating the OR in each analysis.

Additionally, the incidence rate of each outcome of interest was calculated for each trial individually, as well as collectively for participants with and without prior dengue infection. Incidence rates are presented on an annualized basis and per 100,000 persons.

### Definitions for outcomes used in clinical trials

In the Butantan (8) and Takeda (9) studies, the 2009 WHO criteria were used, including cases requiring hospitalization for DVC. In the Sanofi vaccine studies (10–12), DS was defined according to the 1997 WHO definition of dengue hemorrhagic fever, as well as cases requiring hospitalization due to the severity of virologically confirmed infection. An independent study committee also adjudicated cases independently.

## Results

### Identification of Studies and Incidence Rates in Randomized Clinical Trials

A total of 39 publications from three randomized clinical trials were identified. From these, five studies containing the most recent data from the three phase III clinical trials and examining the outcomes of interest were selected (8–12). The PRISMA flowchart is shown in Figure 1, and the risk of bias assessment is presented in Figure 2.

**Figure 1.**
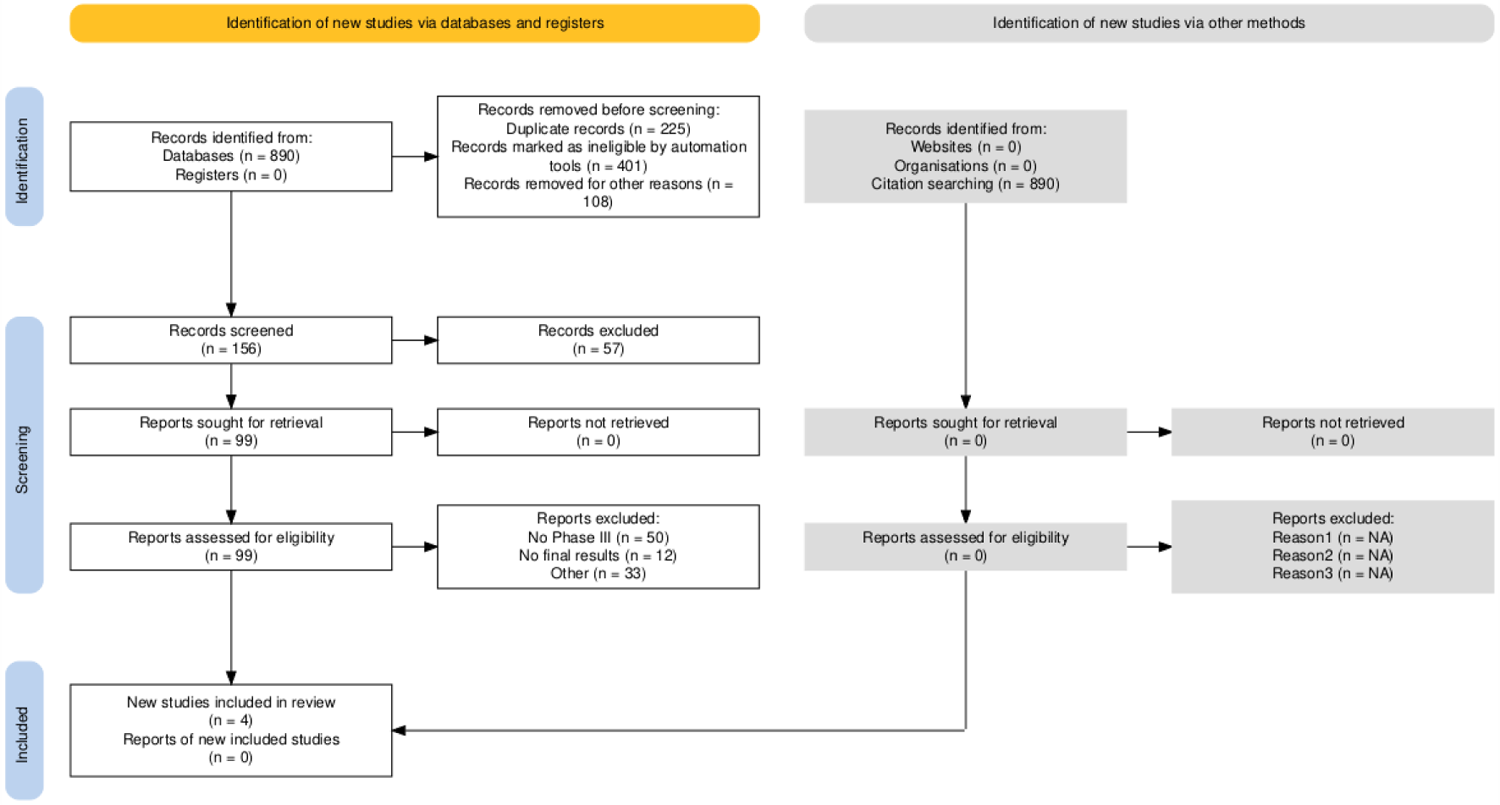
PRISMA flow

**Figure 2.**
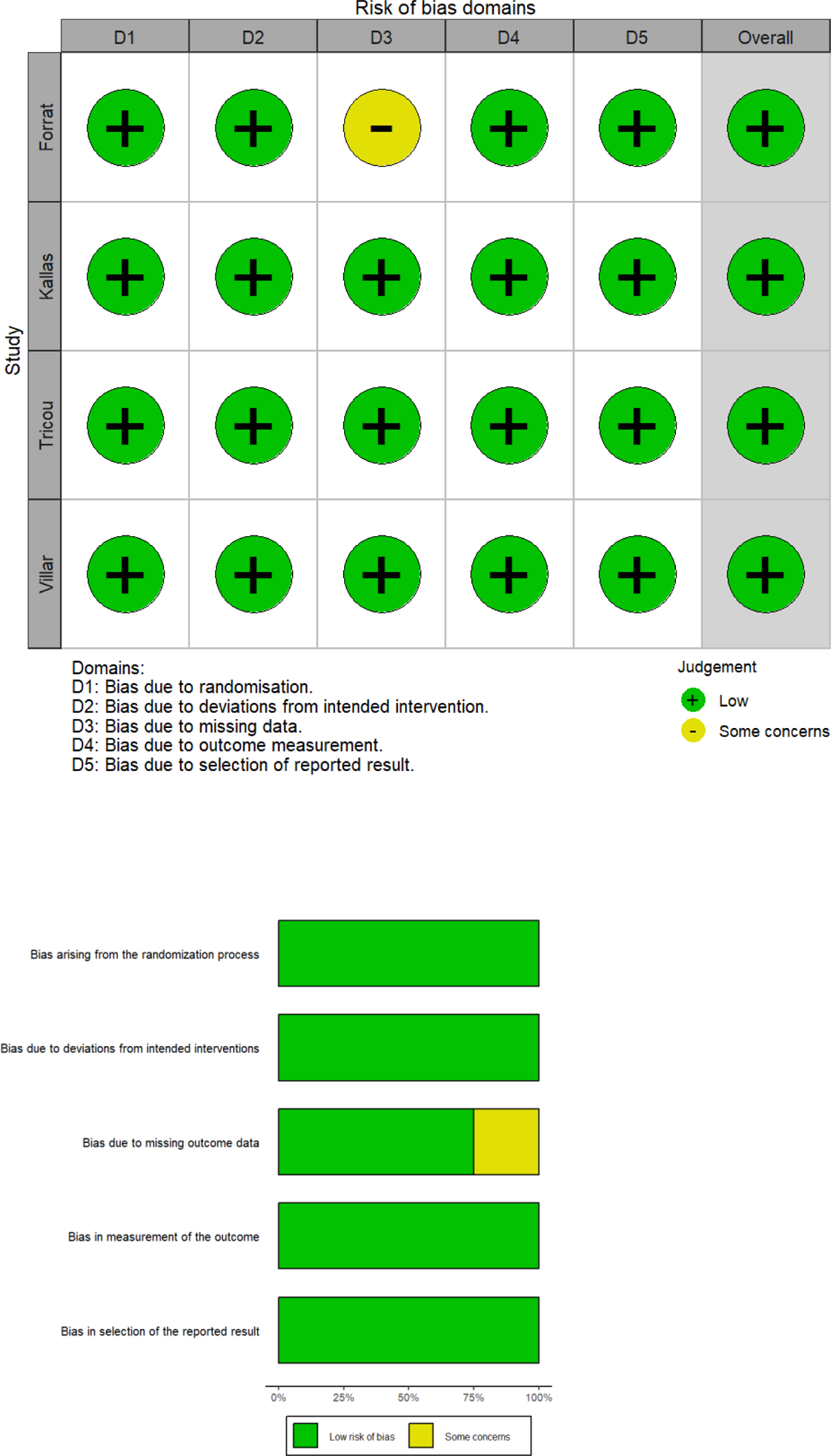
Risk of bias.

### Incidence Rates in Randomized Clinical Trials

The incidence rate of virologically confirmed dengue (DVC) during follow-up was 1,625 (1,536 – 1,715) per 100,000 person-years. The hospitalization rate for dengue was 52 (37 – 68), the rate of severe dengue was 22 (11 – 32), and the all-cause mortality rate was 8 (2 – 14) per 100,000 person-years of follow-up. No dengue-related deaths were recorded in any of the clinical trials during the follow-up periods, which were 6 years for the Sanofi trial (12), 4.5 years for the Takeda trial (9), and 2 years for the Butantan trial (8).

### Association Between Documented Prior Dengue Infection and Outcomes of Interest Virologically Confirmed Dengue (DVC)

Four studies were included with a total of 19,320 observations and 1,084 reported events. Participants with a history of dengue infection had a lower incidence of dengue during the follow-up period, with an odds ratio (OR) of 0.85 (95% CI: [0.75; 0.98], p=0.024) (Figure 3). Heterogeneity among the studies was low, with an I² of 14.5% and a p-value of 0.3197 for the heterogeneity test.

**Figure 3.**
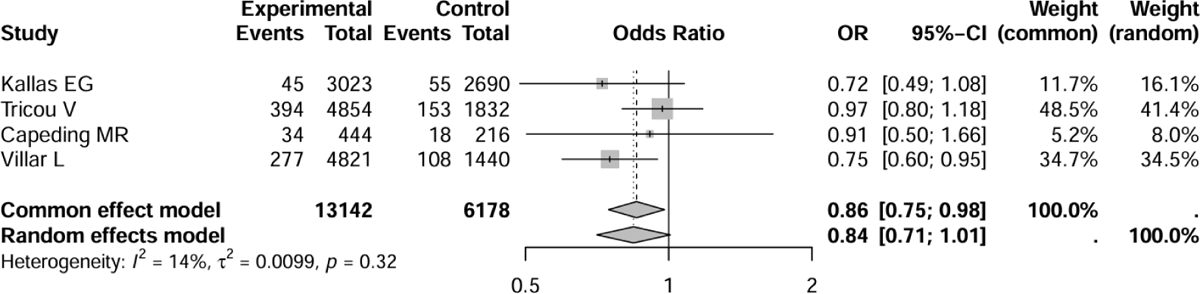
Effect of prior infection with dengue on the incidence of virologically confirmed dengue in the placebo arm of randomised clinical trials

### Severe Dengue (DS)

Four studies were included with a total of 13,493 observations and 57 reported events. The heterogeneity among the studies was very low, with an I² of 0.0% and a p-value of 0.6943 for the heterogeneity test. The fixed-effects model showed that participants with a history of dengue had a higher risk of DS, with an OR of 2.91 (95% CI: [1.23; 6.87], p=0.0149) (Figure 4).

**Figure 4.**
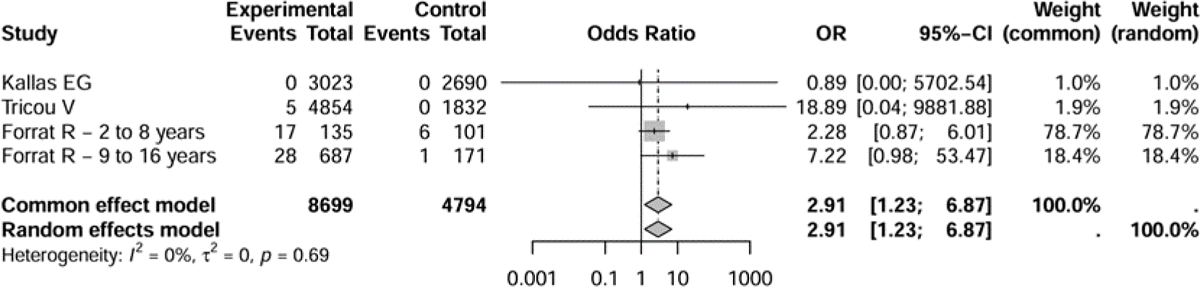
Effect of prior infection with dengue on the incidence of severe dengue in the placebo arm of randomised clinical trials.

### Dengue Hospitalization (DHOSP)

Four studies were included with a total of 13,493 observations and 381 reported events. Participants with a history of dengue had the same likelihood of hospitalization during the follow-up period, with an OR of 1.18 (95% CI: [0.91; 1.53], p=0.198) (Figure 5). The heterogeneity among the studies was moderate, with an I² of 20.1% and a p-value of 0.2893 for the heterogeneity test.

**Figure 5.**
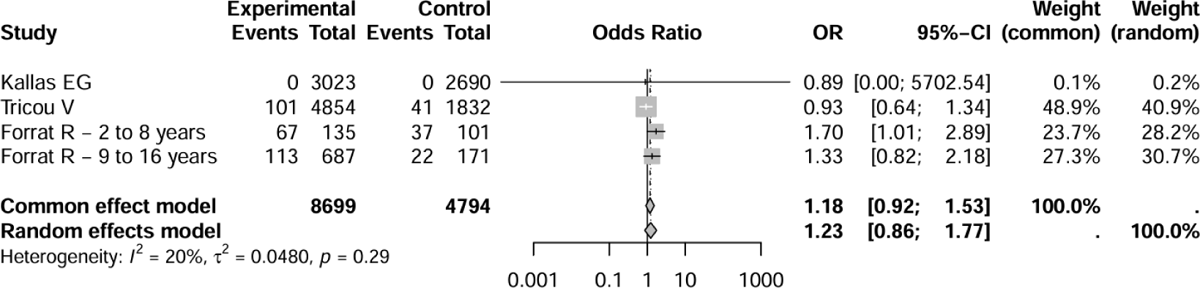
Effect of prior infection with dengue on the incidence of hospitalisation in the placebo arm of randomised clinical trials

### Dengue-Related Death

In the four studies with a total of 13,493 observations, no dengue-related deaths were reported. Since no events were recorded in any of the groups, an adjustment was made by introducing a continuity factor of 0.1. This adjustment allowed for the calculation of an estimated OR and its confidence intervals, which were neutral at 0.50 (95% CI: [0.01; 40.05]).

The presented results did not change significantly when analyzed in a combined form for the two age groups (2 to 8 years and 9 to 16 years) in the Sanofi trials.

## Discussion

This meta-analysis of clinical trials provides valuable information on the severity of events related to a second dengue infection in patients with previously documented dengue. To obtain this information, the frequencies of events were compared in individuals randomized to the placebo arm of vaccine efficacy trials, based on prior seroprevalence.

Patients who were seropositive had a lower risk of secondary infection, similar hospitalization rates, and the same mortality rates as seronegative individuals. However, they were classified as having severe dengue in a higher proportion than those who were seronegative.

The results of this systematic review provide information for a critical reevaluation of the current conceptualization held by the medical community regarding the role of prior infection in the worsening of subsequent infections.

The clinical trials included in this study (8–12) involve all the dengue vaccines approved to date. International health authorities typically make decisions based on the results published in these studies.

The finding of a lower risk of virologically confirmed dengue among participants with previous infections aligns with the well-known fact that prior infection confers homotypic immunity. However, it is described and documented that secondary infection with a different serotype carries a higher risk of severe dengue due to antibody-dependent enhancement (ADE), where preexisting antibodies from the initial infection can facilitate the entry of the virus into host cells, increasing viral replication and disease severity (13). The results reported in this work align with this, showing that the risk of severe dengue was higher in individuals with a prior dengue infection. However, during the follow-up in clinical trials, the most frequent event after virologically confirmed dengue was not severe dengue but hospitalization for dengue. The work reported here shows that the risk of hospitalization for dengue was similar between participants with and without previous disease. This finding has been also published by other groups previously (14–15) and underscores the need to reconsider the utility of the current clinical classification of severe dengue and to reevaluate the role of previous infection as a risk factor for severe events. The results suggest that the definition of severe dengue may not adequately capture the true severity of the disease. On the other hand, the findings highlight the need to adopt hospitalization criteria as a more robust indicator of dengue severity and to reexamine the interpretation of risk associated with previous infections to improve prevention and management strategies for the disease (14–15). Some studies suggest that the revised WHO classification for severe dengue is sensitive and specific (13–15), while other studies indicate inconsistencies and discrepancies in the definition of severe disease (16–20). Although there is consensus that the new WHO classification is simpler and clearer, some studies have pointed out that there may still be difficulties in its application in resource-limited settings (18), as the lack of access to diagnostics and variability in the interpretation of warning signs can complicate its effective implementation (19,20).

In the randomized trials, the number of deaths from dengue during follow-up was zero. This finding was attributed to previous Zika infection in Brazil (21). While this could be biologically plausible for this country, it seems unlikely to explain the absence of deaths in other countries where Zika transmission has been lower or occasional. The absence of deaths from dengue in clinical trials contrasts with the substantial number of observational studies showing distinct lethality (3,7). This contrast can be attributed to several reasons. First, observational studies include patients with different levels of access to medical care and often present higher comorbidity than those recruited in trials. Additionally, patients in randomized studies are closely monitored for symptoms and likely receive earlier medical assistance. This proactive care can prevent the progression to severe forms of the disease and significantly reduce mortality. Second, methodological issues must also be considered. In observational studies, observation bias can lead to an overestimation of dengue mortality risk in patients with previous infections. This bias can arise from several factors, including how participants are selected and monitored, and how data are collected and analyzed. Patients with a history of dengue may be more likely to seek medical care due to awareness of their condition, leading to a higher likelihood of being included in retrospective studies compared to those without a history of dengue.

The work presents limitations that should be considered. First, although only phase III randomized clinical trials were included, the heterogeneity in inclusion criteria and outcome definitions among the studies could have influenced the findings. Second, the limited number of studies and reported events may affect the precision and generalizability of the results. Third, the use of data from the placebo groups of the trials may not fully reflect real-world conditions, as participants in clinical trials often receive more intensive follow-up and superior healthcare compared to the general population. Fourth, the population of the clinical trials included in this meta-analysis consists of those from phase III trials that tested the efficacy of various vaccines. While the efficacy of the vaccine can be extrapolated to other contexts, it should be noted that our work did not assess vaccine efficacy but rather the outcomes of participants in the placebo arm as an indicator of disease burden without vaccination. Thus, the results obtained are indicative of the disease burden in epidemiological contexts with high viral circulation and do not necessarily represent other epidemiological contexts. Finally, the variability in the demographic and geographic characteristics of the studied populations could limit the applicability of the results to other regions and contexts.

In conclusion, the results of this meta-analysis show that prior dengue infection is associated with a higher risk of severe dengue, but not with a higher risk of hospitalization for dengue or increase mortality. Severe dengue in clinical trials represented a smaller proportion of events during follow-up, while hospitalization for dengue was more frequent. These findings suggest that the definition of severe dengue should be revisited to better capture the true severity of the disease, emphasizing hospitalization criteria as a more reliable indicator. The results underscore the necessity of reevaluating the role of previous infections to refine prevention and management strategies for dengue.

## Data Availability

All data produced in the present work are contained in the manuscript

---

**Supplement 1.** Search strategy.

| Database | Keywords | Number of articles |
| --- | --- | --- |
| PubMed | "Dengue fever" OR<br>"Dengue epidemics" AND<br>"Dengue vaccine" OR<br>"Dengue vaccine<br>development" OR<br>"Dengue vaccine<br>prospects" AND "Dengue<br>vaccine efficacy" OR<br>"Dengue vaccine safety"<br>OR "Dengue serotypes" | 890 |
| CINAHL | "Dengue fever" OR<br>"Dengue epidemics" AND<br>"Dengue<br>vaccine" OR "Dengue<br>vaccine development" OR<br>"Dengue<br>vaccine prospects" AND<br>"Dengue vaccine efficacy"<br>OR<br>"Dengue vaccine safety"<br>OR "Dengue serotypes"<br>OR "Clinical | 95 |
|  | trials" OR "Epidemiological studies" |  |
| Cochrane Library | ("Dengue fever" OR "Dengue epidemics") AND ("Dengue vaccine" OR "Dengue vaccine development" OR "Dengue vaccine prospects") AND ("Dengue vaccine efficacy" OR "Dengue vaccine safety" OR "Dengue serotypes") | 72 |
| Web of Science | TS=("Dengue fever" OR "Dengue epidemics") AND TS=("Dengue vaccine" OR "Dengue vaccine development" OR "Dengue vaccine prospects") AND TS=("Dengue vaccine efficacy" OR "Dengue vaccine safety" OR "Dengue serotypes") | 458 |

